# Interventional Neurorehabilitation for Promoting Functional Recovery Post-Craniotomy: A Proof-of-Concept

**DOI:** 10.1101/2021.07.27.21260088

**Authors:** Anujan Poologaindran, Christos Profyris, Isabella M. Young, Nicholas B. Dadario, Syed A. Ahsan, Kassem Chendeb, Robert G. Briggs, Charles Teo, Rafael Romero-Garcia, John Suckling, Michael E. Sughrue

## Abstract

**Purpose:** The human brain is a highly plastic ‘complex’ network –it is highly resilient to damage and capable of self-reorganisation after a large perturbation. Clinically, neurological deficits secondary to iatrogenic injury have very few active treatments. New imaging and stimulation technologies, though, offer promising therapeutic avenues to accelerate post-operative recovery trajectories. In this study, we sought to establish the safety profile for ‘interventional neurorehabilitation’: connectome-based therapeutic brain stimulation to drive cortical reorganisation and promote functional recovery post-craniotomy.

**Methods:** In n=34 glioma patients who experienced post-operative motor or language deficits, we used connectomics to construct single-subject cortical networks. Based on their clinical and connectivity deficit, patients underwent network-specific Transcranial Magnetic Stimulation (TMS) sessions daily over five consecutive days. Patients were then assessed for TMS-related side effects and improvements.

**Results:** 31/34 (91%) patients were successfully recruited and enrolled for TMS treatment within two weeks of glioma surgery. No seizures or serious complications occurred during TMS rehabilitation and one-week post-stimulation. Transient headaches were reported in 4/31 patients but improved after a single session. No neurological worsening was observed while a benefit was noted in 28/31 patients post-TMS. We present two clinical vignettes and a video demonstration of interventional neurorehabilitation.

**Conclusions:** For the first time, we demonstrate the safety profile and ability to recruit, enrol, and complete TMS acutely post-craniotomy in a high seizure risk population. Given the lack of randomisation and controls in this study, prospective randomised sham-controlled stimulation trials are now warranted to establish the efficacy of interventional neurorehabilitation following craniotomy.

## Introduction

The human brain is a highly plastic ‘complex’ network [1,2]: it self-organises without a hard blueprint, it adapts to evolving circumstances, and can withstand external insults. Our thoughts and behaviour are directly governed by how our brain networks handle, orchestrate, and execute various internal and external demands [3]. Nevertheless, similar to other naturally-occurring networks, brain networks can only endure a finite amount of damage before becoming maladaptive and fragmented [4].

The practice of neurosurgery is based on therapeutically altering the brain’s global workspace to improve clinical outcomes [5,6]. However, since the antiquity of neurosurgery, few strategies have been employed to directly address neurological deficits due to iatrogenic injury. In fact, the usual approach is to send patients to physiotherapy and hope they improve over time in a sufficiently stimulating environment. Moreover, rehabilitation is further complicated when surgical pathology implicates critical areas for motor initiation, alertness, motivation, and consciousness [7]. Furthermore, advanced neurocomputational models suggest the capacity for neuroplasticity greatly varies based on the type of cortical damage which has occurred [8]. Ideally, a fundamental goal of neuro-oncological surgery should be to drive cortical reorganisation and promote functional recovery in the immediate post-operative period. To advance this viewpoint, we coin a new concept called ‘interventional neurorehabilitation’: connectome-based therapeutic brain stimulation to promote network plasticity and functional recovery.

Over the past few years, monumental advancements have been made in neuroimaging and neurostimulation technologies. Today, state-of-the art connectome methods enable neuroscientists to make highly accurate single-subject predictions on cognition [9–11]. In addition, we are beginning to non-invasively stimulate focally at-depth without perturbing overlaying cortical structures [12]. However, before leveraging the most advanced technologies for interventional neurorehabilitation, applying well-studied existing stimulation approaches is sensible. Repetitive transcranial magnetic stimulation (rTMS) is an FDA-approved stimulation therapy routinely performed at hospitals across the world [13]. Given its relative ease and non-invasiveness, the field of TMS has flourished to treat a range of neurological and psychiatric illnesses. In acute and chronic stroke patients, rTMS facilitates cortical reorganization leading to functional preservation or compensation in motor and language abilities [14]. Unfortunately, prognosis is still poor in many these patients, which may be explained by the limited capacity for effective cerebral plasticity following some acute injuries compared to slow growing tumors [8]. While meta-analyses highlight the remarkable safety of rTMS in ischemic stroke patients with extremely low-risks for seizures [15,16], there remains limited descriptions on the safety and efficacy of this treatment modality in tumor patients in the acute post-operative period. Given the striking advances in fields outside neuro-oncology, individualised TMS therapy merits investigation to accelerate recovery trajectories post-craniotomy.

In this proof-of-concept study, we sought to establish the safety profile and ability to recruit, enrol, and complete connectome-guided TMS to enhance network plasticity and promote functional recovery following glioma surgery.

## Methods

This study was approved by the Human Research Ethics Committee of the South Eastern Sydney Local Health District (SESLHD). Patients provided written informed consent prior to enrolling in our study. All methods were performed in accordance with relevant guidelines and regulations Declaration of Helsinki. The clinical trial was registered at clinicaltrials.gov (NCT03293888).

### Patient population

Patients with supratentorial gliomas who developed a significant post-operative neurological deficit related to motor or language function were invited to take part in an off-label treatment of FDA-approved rTMS. Subjects were only included in this study if TMS was initiated within two weeks post-surgery. Assessments for motor dysfunction were made using the standard Medical Research Council (MRC) 5-point scale ^[17]^. To be eligible for rTMS therapy, weakness in an arm or leg needed to be 4-/5 or worse in the hand, proximal arm, foot or proximal leg at the time of treatment. Language dysfunction was defined using the Aphasia Rapid Test (ART) with a score greater than 3 considered evidence of significant language disturbance ^[18]^.

### Clinical assessment and Definition of Outcomes

Neurological assessments were performed immediately prior to treatment with rTMS and one week following the last rTMS session by a blinded team member. Improvement in motor function was defined as grade strength to at least 4+/5 in the affected limb, with either functional hand control or the ability to walk with assistance in the leg. In cases of hemiplegia, improvement in either hand or leg function was considered improvement. Finally, reduction in the patient’s pre-treatment ART score by 3 or more points was considered improvement in language.

### Connectome-based TMS target selection in neurosurgical patients

Once recruited, participants underwent a T1-weighted MPRAGE and resting-state fMRI scan. The cortical target was selected based on the patient’s primary deficit (i.e. motor or language), our interpretation of any network fragmentation, and our experience with network topology from normative connectomes (i.e. HCP data) ^[7,19,20]^.

#### Imaging Acquisition and Pre-processing Parameters

The resting-state fMRI was performed on a Phillips 3T Achieva which was acquired as a T2-star EPI sequence, with 3 × 3 × 3-mm voxels, 128 volumes/run, a TE = 27 ms, a TR = 2.8 s, a field of view = 256 mm, a flip angle = 90° and an 8 minute total run time. Resting-state and diffusion pre-processing was performed using in-house custom machine learning algorithms in Python. Standard image processing steps included skull stripping, motion correction with a 6-dimensional rigid body registration, correcting for physiological noise (CompCor), slice time correction, spatial smoothing (6 FWHM Gaussian kernel), high-pass filtering, and co-registration to the patient’s structural space^1^. Of critical importance, we do not warp the brain into a standard space like Montreal Neurological Institute (MNI) or Talairach space at any stage of the processing. The patients then had a diffusion sequence acquired for subsequent connectivity analyses from patient-specific multi-modal imaging data.

### Machine-learning Aided Parcellation for Brain Tumours

A fundamental challenge for interventional neurorehabilitation post-craniotomy is to apply a parcellation scheme to highly-distorted anatomical brains. The Glasser HCP parcellation scheme is a state-of-the art multi-modal neurobiological division of the cerebral cortex ^[21]^. However, it was not designed to be applied to brains with large lesions and oedema. We aimed to directly address this challenge by determining new HCP parcellation locations by using a proprietary machine learning algorithm (Omniscient Technologies) –Figure 1 is the connectome construction pipeline and Figure 2 represents sample outputs. Using a supervised machine learning approach, we first trained our algorithms to identify each HCP parcel using network connectivity from a normative dataset. Then, we applied our machine to identify the most appropriate HCP parcels in brains after supratentorial tumour surgery based on the same input imaging data. To our knowledge, this approach is unique in that previous studies have resolved this issue by applying the HCP parcellation derived from healthy brains without any adjustment to cortical topology.

**Figure 1:**
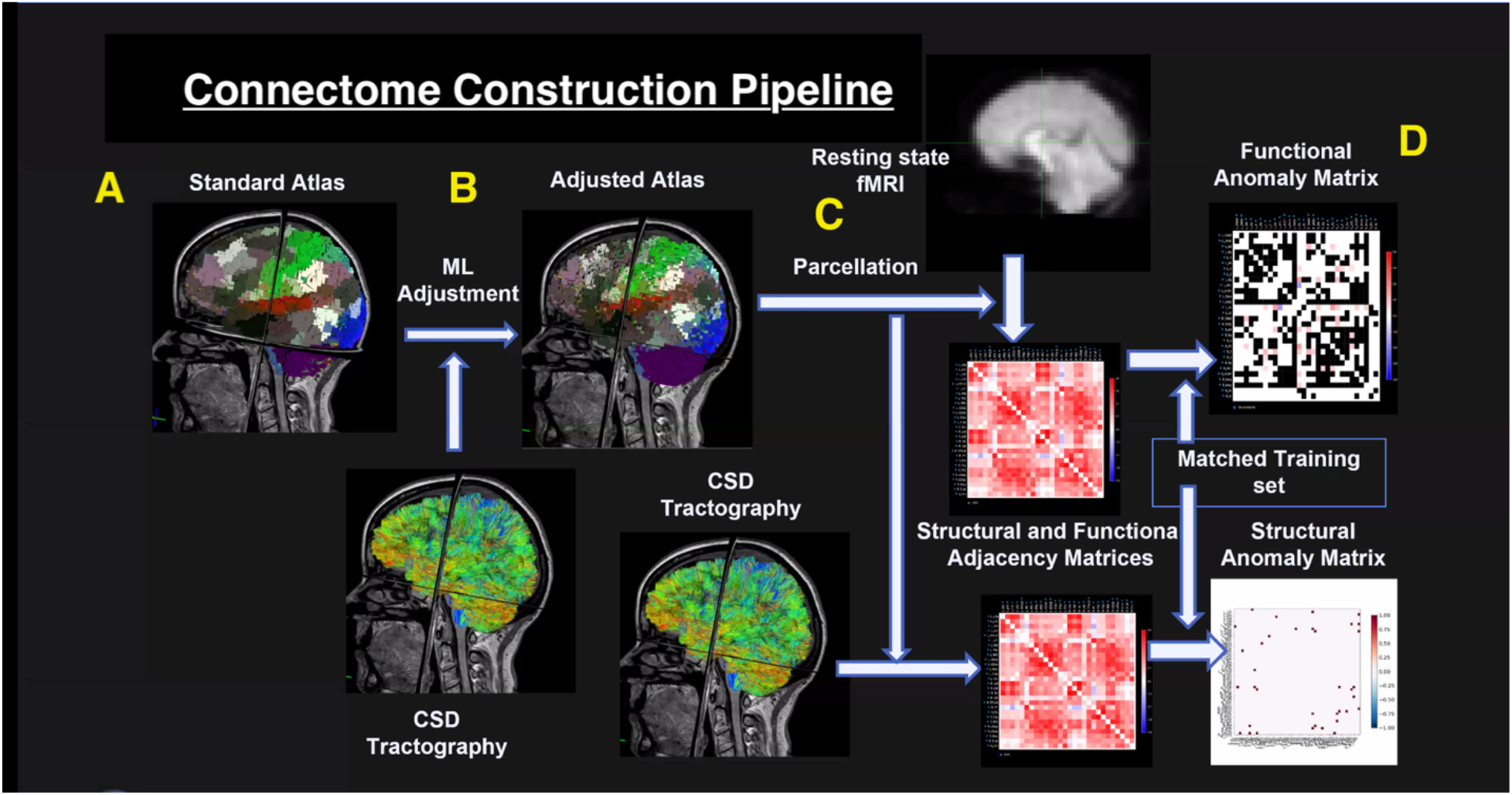
The connectome construction pipeline used in this study. A) A standard Glasser atlas was established using 300 healthy individuals from the Human Connectome Project (HCP). A supervised machine learning algorithm was employed to recognise connectivity patterns for each of the 360 HCP parcels in a healthy cohort. B) Using diffusion sequences, we applied constrained spherical deconvolution (CSD) tractography to our patient cohort. Using these images, our algorithm was applied to recognise and adjust the locations of HCP parcels in highly atypical brains. C) After establishing maximal likely structural connectivity, we used this data to inform and constrain functional connectivity using resting-state fMRI. D) Finally, structural and functional anomaly matrices were generated to compare network connectivity differences (i.e. language) between our patient and a normative atlas. Adopted with permission from Reference ^[48]^.

**Figure 2:**
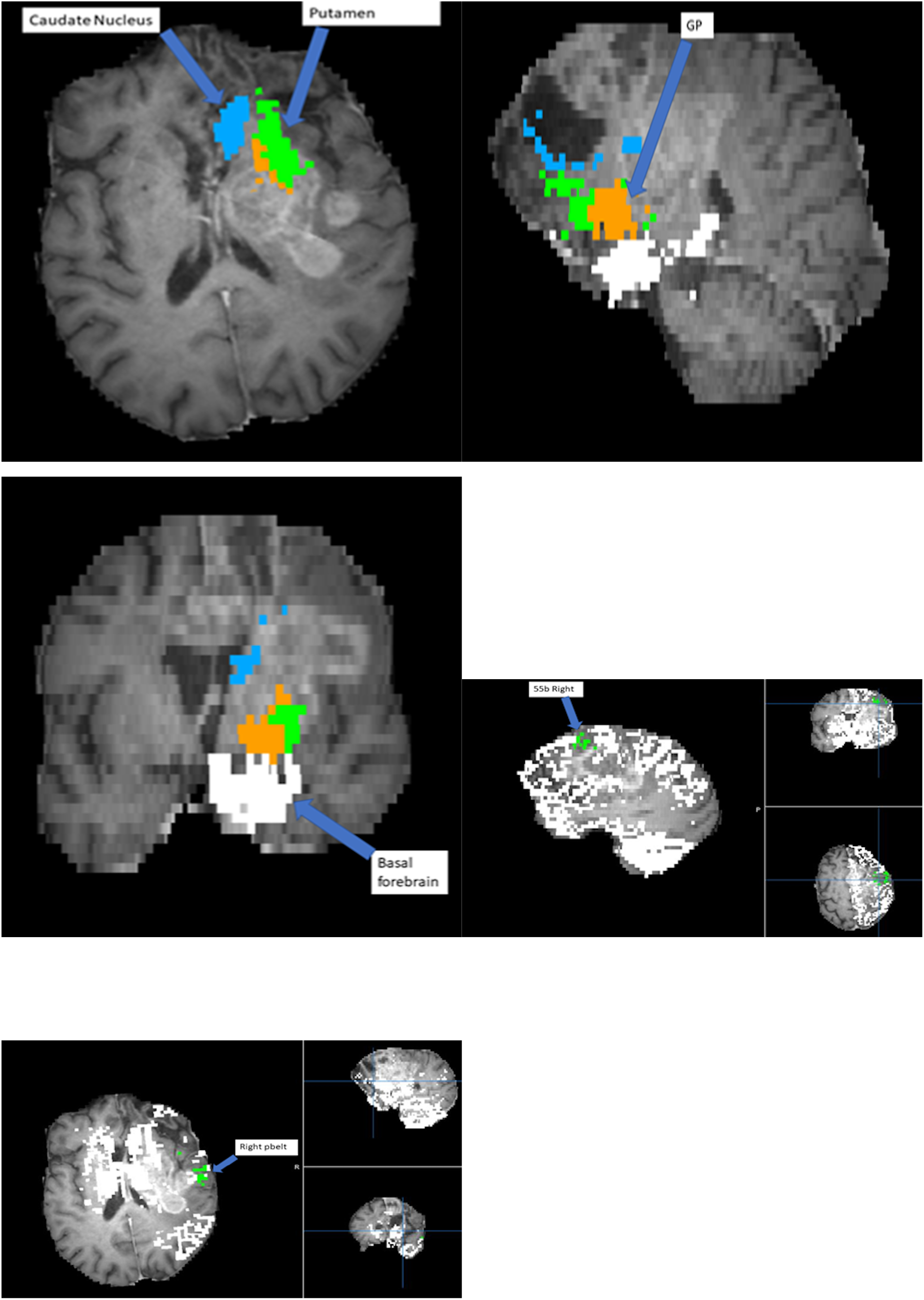
Demonstration of proprietary machine learning algorithm (Omniscient) that assigns parcellations to very distorted brains. Patient with a frontal lobe GBM and resected regions resulting in total anterior brain shift. Figure 2a displays the modified location of the caudate nucleus and the putamen. Figure 2b displays the modified location of the GP. Figure 2c displays the modified location of the basal forebrain. Figure 2d displays the modified location of right 55b parcellation. Figure 2e displays the modified location of the right PBelt. This allows for the creation of a connectivity matrix of any brain despite.

### Comparative Connectome Analyses

To gain additional insight into network connectivity, we processed n=300 HCP connectomes to serve as a reference of healthy canonical brain network organisation. Using this normative data, we qualitatively compared healthy networks to those observed in patients with lesions in particular areas. For example, we compared the normative visual areas to a patient with hemianopia (Figure 3a) or normative language network topology with that of a patient with aphasia (Figure 3b). This intra-network analysis enabled us to perform a hypothesis-driven neuro-navigated rTMS target selection.

**Figure 3:**
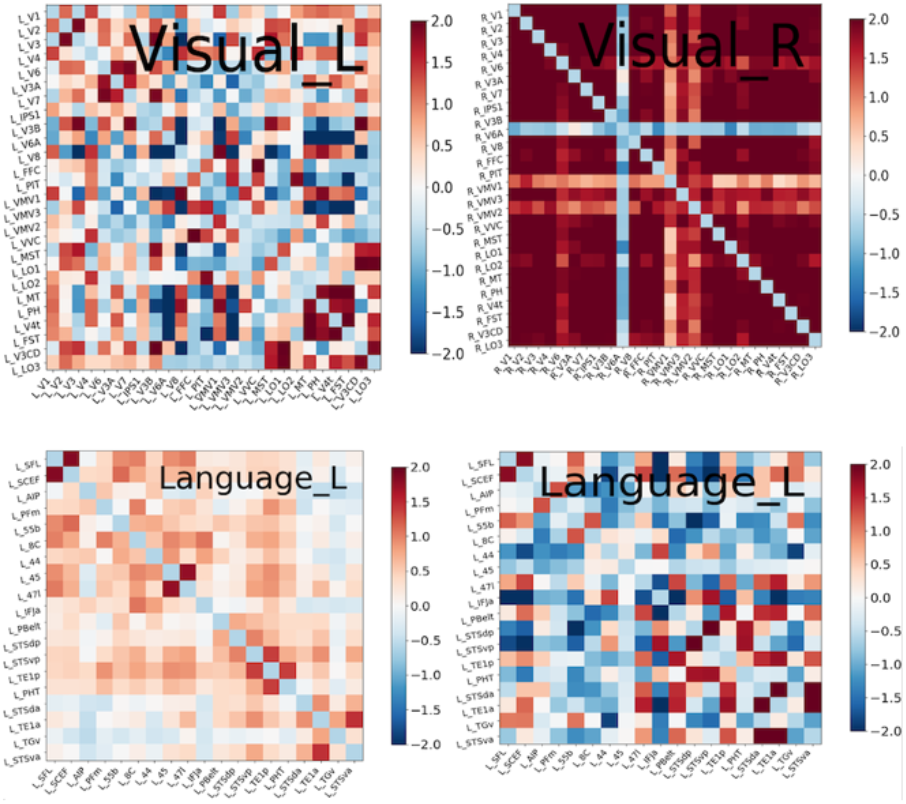
3a compares connectivity matrices of the left and right visual networks in a patient with hemianopia. The left visual network is dotted mostly *blue*, which means that areas of the visual system are not well synchronized to one another. By comparison, the right visual network displays strong intra-network connectivity. Figure 3b compares the connectivity matrix of the language area of a healthy control on the left with the language area connectivity of an aphasic patient on the right. This aphasic matrix has the parcellations within the language system anticorrelated, therefore, predominantly *blue*, suggestion loss of connectivity within the language network. Note that columns 55b, 45 and STSdp are blue representing that they are isolated. We hypothesized that this is in part due to problems with the superior longitudinal fasciculus/ arcuate fasciculus system which links different components of the language system^7^. Conducting connectomics analysis by comparing connectivity matrices enables us to generate potential targets for TMS treatment.

### rTMS treatment paradigm

The rTMS treatment was initiated within 1-2 weeks after standard awake glioma surgery. We utilized theta burst stimulation (TBS) protocols in all patients. Details of the TMS protocol and rationale available in the SI. We performed treatment five times per day over five consecutive days. In between TMS sessions, patients underwent rehabilitative therapy.

### Complications and Adverse Events

All complications and side effects were noted after each rTMS session and one-week post-treatment. Seizures were defined as any observable seizure or possible seizure-like activity during the course of treatment. Neurological complications included any new or worsening of neurological dysfunction measured by the ART and MRC Motor scale.

## Results

### Preliminary safety and recruitment data regarding rTMS treatment in neurosurgical patients

We successfully recruited 31/34 (91%) patients within two-weeks after glioma surgery and treated them with rTMS. The median participant age was 58 years with 20 females and 14 males. 30 patients had WHO grade II-IV gliomas, while four patients had low grade gliomas. Of all the participants, n=23 began rTMS therapy within a week of surgery, and n=31 began within 2 weeks of surgery. The remaining 3 participants underwent treatment at 2 months, 4 months and 12 months and excluded in the recruitment rate citing logistic concerns. In total, 31 participants completed all planned treatment sessions with one participant missing one rTMS session due to a rehabilitation bed becoming available the day of their last scheduled treatment. No participant stopped therapy due to treatment intolerance. In 21 participants with a motor deficit, rTMS was applied to the sensorimotor network with an improvement noted in 19 patients after one-week following the last TMS session. In 13 participants with a language deficit, rTMS was applied to the frontoparietal network with an improvement in 12 patients after one-week of the final stimulation session.

#### Safety and preliminary efficacy of rTMS in neurosurgical patients

No participants reported any general or partial seizures or seizure-like events during the course of treatment and follow-up. Four patients reported transient headaches which resolved at the end of each individual session. Light headedness (n=1) and nausea (n=1) was also reported but resolved before the start of the next session. Transient tingling was reported at the site of stimulation during stimulation onset, but also resolved immediately. These results are consistent with well-documented side-effects during rTMS of non-craniotomy patients ^[13,16,22]^. We noted no worsening of neurological deficits and no other obvious side effects. The Supplemental Digital Content is a video of a typical procedure; the participants consented to publication of his/her image. Two brief clinical vignettes are presented below.

### Clinical Vignettes

#### Case 1

A woman (age 60-65) with a left parietal glioblastoma presented with preoperative aphasia and near complete hemiplegia. Following resection, she developed complete expressive aphasia and right hemiplegia. Connectome analyses revealed that her sensorimotor networks were fragmented as two independent parcellations, likely due to the destruction of the callosal fibers. Specifically, the injured side demonstrated satellite areas anterior to the dysfunctional sensorimotor networks (Figure 4). Additionally, the left frontoparietal network revealed a clear component of Broca’s area, area 55b, and an SMA component. However, the temporal component appeared to be less organized, appearing abnormal compared to normative data. Thus, to potentially enhance functional recovery and address both delocalised networks, we sought to select a stimulation target that would lead to enhanced network recruitment.

**Figure 4:**
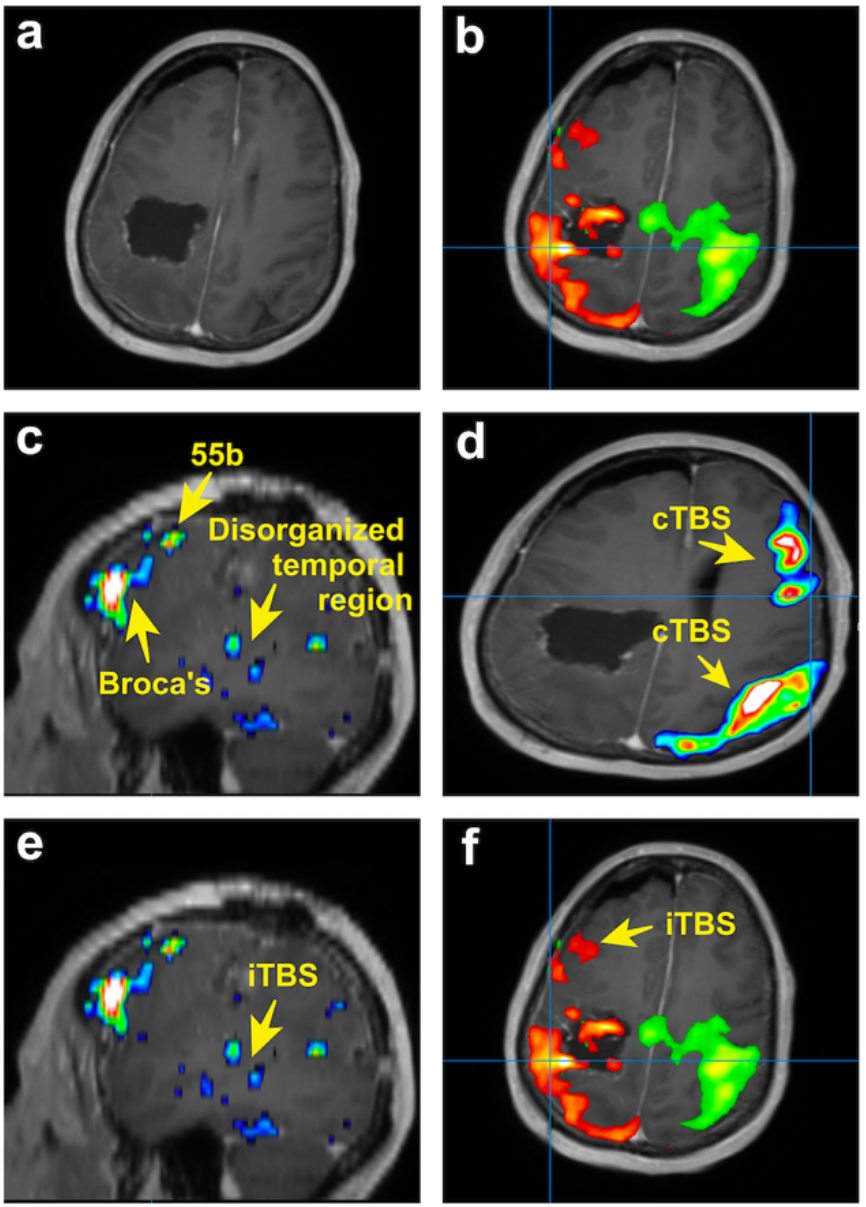
TMS strategy for patient presenting with aphasia and near complete hemiplegia secondary to glioblastoma. (a) Postoperative MRI of patient demonstrating resection cavity. (b) Independent right sided (green) and left sided (orange) sensorimotor networks. Although presented on the same image, these networks appeared as separate networks on connectomic analysis. The anterior satellite areas in the left (orange) dysfunctional sensorimotor network. (c) Left frontoparietal network demonstrating clear Broca’s area and area 55b. The temporal component of the network is disorganized. (d) cTBS was administered to both the middle of the sensorimotor network and the right posterior frontal component of the right frontoparietal network. (e) iTBS was administered to the disorganized temporal component of the left frontoparietal network and the (f) anterior areas of the pathological left sensorimotor network.

Beginning on post-operative day five, we performed five days of daily continuous TBS (cTBS) to both the middle of the right sensorimotor network and the posterior frontal component of the right frontoparietal network (both targets treated once per day). We then performed intermittent TBS (iTBS) to the areas of scattered activation in the posterior left temporal lobe and the areas near the abnormal sensorimotor regions. This treatment was well tolerated, and by the end of the treatment, the patient was able to ambulate with a cane and speak in full sentences. There were no serious complications, however, she had some persistent arm weakness.

#### Case 2

A man (70-75 years of age) with a posterior left insular glioblastoma had moderate pre-operative expressive aphasia that persisted post-surgery. Connectome analyses demonstrated that his posterior temporal region was appropriately organized but did not co-activate within the same network as Broca’s area (Figure 5) [6]. Thus, we hypothesized that this was the result of inactivation of the arcuate fasciculus fibers by the tumor or related to oedema. We also noted that he was recruiting the right analog of Broca’s area, as both regions were functionally co-activated. As a result, we chose to perform accelerated (spaced-delivery of stimulation ^[23]^) iTBS to the left posterior temporal site to enhance the recruitment of additional connections for speech improvement. This treatment began on post-operative day four. At the end day five of rTMS, his speech markedly improved with no complications to report. Nevertheless, he persisted with residual paraphasia after his therapy.

**Figure 5:**
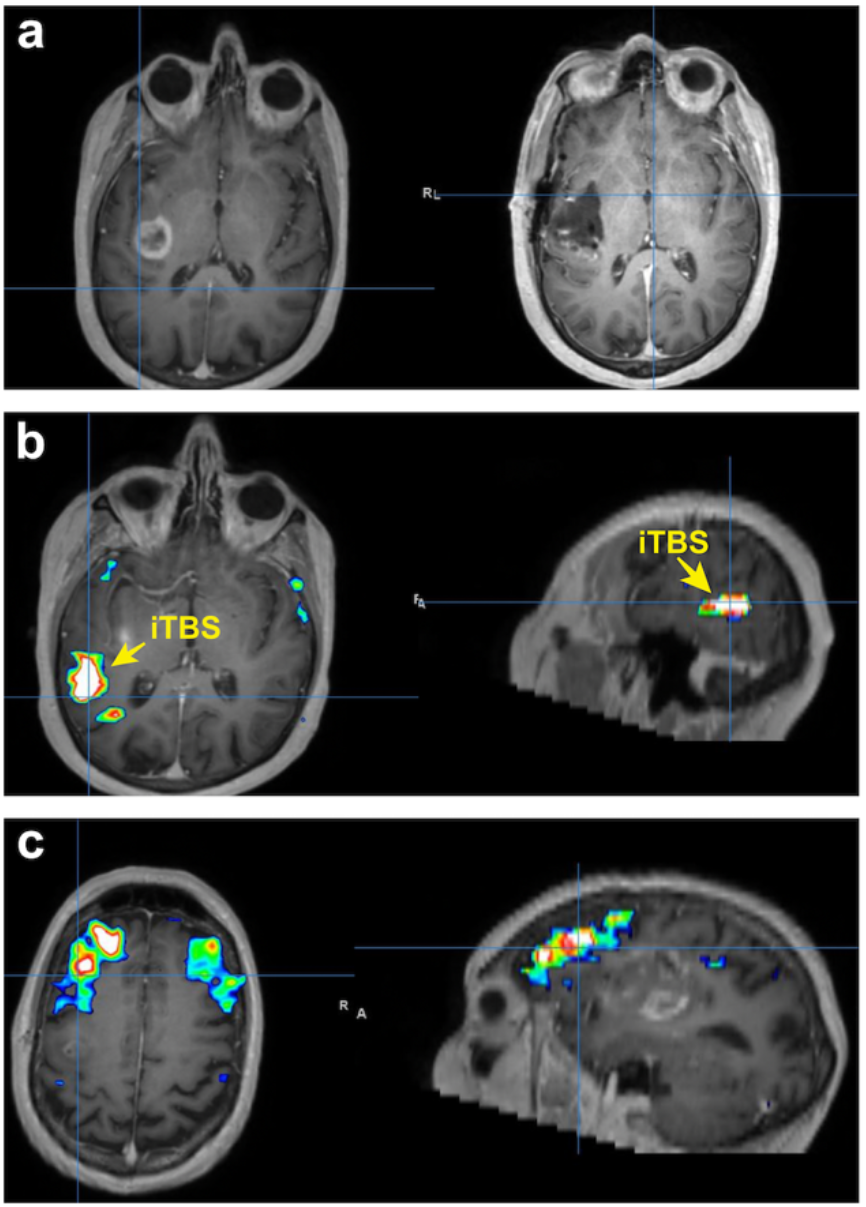
TMS strategy for patient presenting with moderate expressive aphasia secondary to glioblastoma. (a) Preoperative MRI (left) demonstrating left insula glioblastoma and postoperative MRI (right) demonstrating complete resection. (b) Network analysis demonstrating a strongly organized posterior temporal region that is not in in the same network as Broca’s area. This was the area that was selected for treatment with iTBS. (c). Further network analysis demonstrating Broca’s area with bilateral representation that is not in the same network as the posterior temporal region.

## Discussion

In this study, we demonstrate the safety of rTMS post-craniotomy with the goal of promoting functional recovery. Specifically, we demonstrate that no seizures were induced in 31 patients post-craniotomy and transient side effects were reported in 6 patients. This work complements safety data from dozens of rTMS studies completed in non-craniotomy individuals ^[24,25]^. Despite the uncontrolled and open-label nature of the study, we cautiously interpret that rTMS can potentially facilitate functional recovery post-craniotomy. Similar results have been illustrated in acute and chronic stroke patients suggesting the possible role of TMS as a therapeutic modality for a variety of clinical conditions to facilitate motor and language improvement ^[14]^. Given the widely demonstrated safety profile of TMS, it would be a disservice not to further investigate the efficacy of technology to optimize post-surgical clinical outcomes. To fully harness interventional neurorehabilitation’s potential for neuro-oncological care, additional research is required in two areas: target engagement and simulation protocol. Here, we elucidate the role of individualized TMS in standard inpatient rehabilitation and discuss implications for future study on rTMS to optimize clinical outcomes.

### Importance of Target Engagement and Stimulation Protocol

There is a growing body of evidence suggesting that effective TMS targeting is critical for success. For example, using image guidance to target rTMS improves efficacy ^[26]^. Furthermore, targeting brain networks affected by disease-related processes is crucial for functional improvement. Recently, Momi and colleagues delivered TMS pulses to two frontoparietal nodes (prefrontal and parietal) to enhance fluid intelligence tasks ^[22]^ – adding another research consideration on multi-nodal, rather than uni-nodal, stimulation. In addition, it is likely that different patients with the same clinical deficit may need different target(s) ^[27]^. Hence, there are many ways to interpret these observations, however, we advocate establishing “the right target for the right patient” as being critical to successful interventional rehabilitation.

Similar to target engagement, *stimulation protocol* is another important variable to consider. There are numerous different TMS protocols available for use. However, TBS protocols are better suited for neurosurgical patients. First, the lower stimulus intensities used in TBS likely have a lower seizure risk ^[28]^. Second, TBS protocols achieve similar effects with shorter treatment times (typically 8 minutes per session) compared to standard 30 minutes with 10 Hz TMS protocols. This enables the use of accelerated protocols (spaced-delivery of stimulation sessions ^[23]^) which are useful in treating patients in a subacute paradigm. Finally, the stimulation effects of TBS is believed to last 45-60 mins which may fit better when coordinating inter-session rehabilitation ^[29,30]^. Our view is that while seizures are a concern, given our clinical experience in managing this problem concomitant with the low occurrence rate of this complication, there is now sufficient evidence to justify offering neurosurgical patients rTMS.

#### Connectome-based Stimulation for Cognitive Rehabilitation

In this study, we primarily focused on ameliorating motor and language deficits post-surgery in glioma patients. However, many patients experience cognitive deficits post-surgery and there are no clear guidelines on how to help these patients. The multiple demand (MD) system is a domain-general cognitive control network that acts as a skeleton for executing cognitive tasks [3,7]. Systematically studying this system and the implications of its removal during surgery would be useful for predicting post-operative cognitive trajectories ^[31]^. More broadly, despite motor or language deficits in our cohort, the qualitative fundamental motivation to rehab greatly varied between our individuals ^[32]^. An increasing line of evidence suggests that increasing the motivation to expend cognitive effort, rather than enhancing cognitive networks themselves, would be more effective in bolstering goal-directed behaviour ^[33]^. Thus, if frontostriatal circuitry can be mapped and effectively modulated post-craniotomy, this would be a significant advancement and become important for other areas of neurosurgery, such as limbic surgery ^[34]^.

### Establishing a TMS Clinic for a Neurosurgical Practice

While rTMS is a well-developed field with standard techniques, treating post-craniotomy patients posed some unique challenges. First, the benefit of TMS early in the post-operative period appears important ^[35]^. Due to logistical issues, not all patients were able to return within one week of surgery and returned within 14 days. However, there are important limits to early TMS intervention in neurosurgical patients. Chiefly, given the small risk of inducing a seizure, patients with significant swelling and/or midline shifts are not optimal candidates for rTMS. Furthermore, patients with seizure history due to a tumour should be excluded from TMS until more evidence of its safety in these circumstances are available. Despite these issues, we now feel rTMS can be safely performed as part of standard inpatient rehabilitation.

### rTMS Therapy for Stroke and Surgery

The literature on the role of TMS in motor and language functional recovery is well-established in stroke patients, providing most of our insight into the current benefits and limitations of this therapeutic modality. Thus, certain themes from stroke neurology may cautiously applied to neuro-oncological patients to guide therapeutic stimulation. For example, cerebral inflammation and angiogenesis are two of multiple overlapping processes between glioma surgery and cerebral ischemic stroke pathways [36]. A recent meta-analysis on 841 patients across 20 randomized controlled trials (RCTs) demonstrates that rTMS is beneficial to the treatment of post-stroke hemiplegia, especially in: lower limb functioning, grip strength, and attenuating stroke severity [15]. Interestingly, cortical reorganization can be observed between primary motor and secondary motor cortices in stroke patients to facilitate improved motor functioning, leading authors to suggest the need for future customized TMS applications based on the newly activated cortical pathways in these stroke patients [37]. Similar cortical re-organization has also been demonstrated to facilitate language functioning following ischemic stroke [38]. Thus, following iatrogenic injury due to tumor resection, it is likely that similar cortical pathways responsible for motor functioning and motor learning can also be strengthened with rTMS given they too demonstrate plasticity following tumor growth [39,40].

rTMS likely provides additional benefits in many patients following tumor resection, while being precluded from some stroke patients, due to certain temporal factors which are known to affect plasticity. For instance, certain gliomas may cause slower changes in cerebral connections and therefore affect connectome plasticity differently compared to acutely damaging stroke lesions[39]. Unfortunately, acute brain damage due to stroke often causes more localized neuronal cell death and subsequent unimodular cortical organization [8]. This type of damage may demonstrate less capacity for cerebral reorganisation compared to slow growing tumors (ie, low-grade gliomas, LGG), which disrupts greater amounts of cortex but provides more opportunity for functional reorganization possibly due to less abrupt, initial neuronal death [8]. Thus, even more importantly in tumor patients, individualized connectomic approaches can be implemented before rTMS to identify any on-going network reorganization occurring from the lesion and ultimately facilitate motor and language preservation or compensation. This would enable an updated understanding of network connectivity which can be applied to effectively target and strengthen formerly silent polysynaptic cortical pathways following tumor resection ^[14,40]^. Indeed, our study shows that intra-network analyses can *safely* enable hypothesis-driven neuro-navigated rTMS target selections.

While the applications of TMS between stroke and surgical patients have not been compared, their respective methods may differ and therefore preclude certain benefits which have been demonstrated specifically in ischemic stroke patients. Unfortunately, no studies were able to be identified which looked at post-surgical rehabilitation utilizing individualized rTMS treatment. Most surgical studies incorporating TMS have confined their uses to intra-operative functional mapping (ie, language mapping) [41]. The only post-surgical application of rTMS identified investigated its ability to reduce pain following gastric bypass surgery [42]. Nonetheless, in stroke patients, the effects produced by TMS correlates with the specific TMS protocol applied ^[14]^. Specifically, a TBS protocol of 50 Hz with 80% of active motor threshold intensity is believed to modulate motor learning through effects similar long-term potentiation (LTP) and long-term depression (LTD), and has been applied in numerous stroke trials ^[14,43]^. Our study utilized bursts of 3 pulses delivered with a repetition rate of 50 Hz at 80% of motor threshold for 400 continuous (cTBS) or intermittent (iTBS) trains. Still, while our protocol is similar to those described for ischemic stroke patients and we too observed improvements in language and motor functioning, possible differences in neuroplasticity as identified by neurocomputational models between glioma and stroke patients suggests that specific rTMS protocols may produce different cortical changes and subsequent functional recovery based on the type cortical injury.

### Future Directions for Interventional Neurorehabilitation

With the rapid advancements made in imaging and stimulation technologies [32], the future is bright to continuously evolve the concept of interventional neurorehabilitation for neuro-oncology. Importantly, pre-operatively predicting which patients will require post-operative interventional support or predicting which patients will best respond to and benefit from rTMS as a therapeutic adjunct would be highly valuable. rTMS treatment causes changes in electroencephalographic (EEG) assessed-functional connectivity (FC) that correlates well with clinical outcomes in certain patients. Thus, EEG-FC measures can provide accurate, early, biomarkers to guide personalized clinical decisions for long-term rTMS treatments or not, possibly saving hospital costs while improving patient quality of life over time [44]. A machine learning algorithm can accurately predict (83%) which patients with major depressive disorder (MDD) will respond to long-term rTMS treatment in just the first rTMS session [45]. Similar studies have not been extended into the perioperative setting, but numerous studies have established the role of TMS and fMRI data as biomarkers to predict motor recovery following stroke, warranting further investigation following neuro-oncological resection [46,47]. Lastly, interventional neurorehabilitation can also be utilised in other patient populations following stroke-induced pain or movement disorders [48–50]

With rising open science and international collaborative efforts, prospective registry-based clinical studies could be employed to overcome heterogeneous glioma populations and derive more practical outcomes.

### Study Limitations

This study, however, is not without key limitations. First, the uncontrolled and early-nature of our intervention raises the possibility whether these patients would have improved without treatment. Now that we have established the TMS safety profile and recruitment rate for this complex patient population, future trials should employ prospective, randomised, double-blinded active- and sham-controlled TMS to determine the efficacy of improving recovery trajectories. While glioma patients are highly heterogenous given different tumors, different degrees of cortical reorganisation, and different resections completed, strict inclusion criteria and multi-site collaborations can overcome such limitations. An alternative approach would be to conduct large-scale international prospective registry-based studies. Finally, we did not acquire long-term outcome data in these patients. Nevertheless, qualitatively, all patients reported they would undergo TMS therapy again and found that it assisted with their rehabilitative efforts. Despite these limitations, our primary aim was to establish the safety profile and recruitment rate for TMS post-craniotomy.

## Conclusion

In conclusion, we present a proof-of-concept of ‘interventional neurorehabilitation’ for neuro-oncological clinicians to take charge in driving cortical reorganisation and functional recovery. Specifically, we demonstrate the safety profile and recruitment rate for connectome-based TMS acutely post-surgery for glioma patients. Given the clear enthusiasm from our patients, we believe that TMS treatment is of low-risk, well-tolerated, and could be of immense therapeutic benefit.

## Data Availability

Data available upon reasonable request and clinical ethics approval.

## Acknowledgements

AP is funded through a Turing Doctoral Scholarship from the Alan Turing Institute and the National Science and Engineering Research Council of Canada. RRG is supported by a *Guarantors of Brain* Fellowship.

## Financial Disclosures

IMY is an employee of Cingulum Health. MES is the chief medical officer of Omniscient and a shareholder of Cingulum Health. CT is a consultant for Aesculap. All other authors report no conflict of interest related to this study.

## Author Contributions

Conceptualization, AP, IMY and MES; Data curation, CP, IMY; Formal analysis, AP, IMY.; Methodology, AP, RGB, MES; Project administration, IMY, CT. and M.E.S.; Software SAA, KC, RRG; Writing—original draft, AP.; Writing—review and editing, NBD, RRG, JS, MES.

## Supplemental Digital Content

This video expands on the TMS technique described in the Methods. The video illustrates patient set-up, patient registration, measuring motor threshold, and TMS treatment. The participants consented to publication of his/her image. https://www.dropbox.com/s/prp7dm3h7bsgm62/TMS_Demonstration_Video.mp4?dl=0

## Notes

### Clinical Trial

NCT03293888

### Funding Statement

AP is funded by a Turing Doctoral Scholarship from the Alan Turing Institute and the National Science and Engineering Research Council of Canada. RRG is supported by a Guarantors of Brain

### Author Declarations

Human Research Ethics Committee of the South Eastern Sydney Local Health District (SESLHD)

